# Altitude and Delayed Cerebral Ischemia After Aneurysmal Subarachnoid Hemorrhage on the Tibetan Plateau: A Retrospective Cohort Study

**DOI:** 10.64898/2026.06.19.26356110

**Authors:** Zhenbo Song, Chunmei Hu, Danbai Wujin, Yujie DuoJi, Xubin Chang, Xudong Cao, Zeng Ren, Guangyong Wu

## Abstract

**Background:** Delayed cerebral ischemia (DCI) is a major driver of poor functional outcome after aneurysmal subarachnoid hemorrhage (aSAH). Virtually all DCI risk factor evidence derives from sea-level populations, with few data examining DCI as a primary endpoint in patients chronically exposed to high-altitude hypoxia.

**Methods:** This single-center retrospective cohort study at the People’s Hospital of Tibet Autonomous Region included 256 consecutive aSAH patients residing at 2330–4920 m. DCI was adjudicated per 2010 international consensus criteria, with residential altitude and admission hemoglobin prespecified as primary exposures. A prespecified multivariable logistic model (Model A; events-per-variable ≥10) identified independent DCI correlates, with sensitivity analyses across three DCI definitions and internal validation via 2000 bootstrap resamples with calibration assessment.

**Results:** DCI occurred in 60/256 patients (23.4%). Fisher grade III–IV (aOR 3.38, 95% CI 1.77–6.47) and surgical treatment (aOR 5.22, 95% CI 2.37–11.47) independently predicted DCI. Each 100-m residential altitude increment associated with higher DCI odds but fell just short of significance (aOR 1.08, 95% CI 1.00–1.17; p=0.051), with unstable significance across model specifications (p=0.021–0.157). Admission hemoglobin showed no association (aOR 1.00; p=0.988). Model A demonstrated acceptable discrimination (AUC 0.759, bootstrap 95% CI 0.690–0.828). DCI strongly associated with increased in-hospital mortality (20.0% vs 3.6%) and 1-year functional dependence (mRS ≥3: 47.9% vs 14.8%).

**Conclusions:** In this high-altitude aSAH cohort, hemorrhage burden and surgical treatment were the dominant DCI correlates, while admission hemoglobin showed no association. Residential altitude demonstrated a suggestive borderline DCI signal most pronounced above ∼4000 m, consistent with a potential threshold rather than linear effect. These hypothesis-generating findings require multicenter prospective validation before clinical application.

## 1. Introduction

Aneurysmal subarachnoid hemorrhage (aSAH) represents a small proportion of all stroke cases yet contributes disproportionately to stroke-related mortality and years of potential life lost, largely because it strikes younger patients and carries substantial early case fatality [1]. Among the secondary complications that determine long-term functional outcome, delayed cerebral ischemia (DCI) stands out as the leading potentially preventable cause of death and disability following the initial hemorrhage [2]. DCI complicates roughly 30% of aSAH cases, typically developing between 4 and 14 days after ictus through a multifactorial process involving large-artery vasospasm, microcirculatory dysfunction, microthrombosis, and cortical spreading depolarization. It is strongly linked to cerebral infarction, prolonged intensive care unit stays, and poor functional recovery [2]. Recognizing its prognostic significance, a multidisciplinary research group established a consensus definition of DCI to standardize its use as an outcome endpoint in both clinical trials and observational studies [3]. Despite advances in aneurysm securing techniques and neurocritical care, DCI incidence has declined only modestly, and reliable strategies to predict and prevent it remain an important unmet clinical need.

A large body of work has delineated the principal determinants of DCI in conventional, low-altitude populations. Systematic reviews and meta-analyses consistently identify high Hunt-Hess and World Federation of Neurosurgical Societies (WFNS) grades, thick cisternal blood on admission computed tomography (reflected by high modified Fisher grade), female sex, hypertension, and smoking as independent predictors [4,5]. Building on these established factors, several bedside risk stratification tools have been developed, including the VASOGRADE, which combines WFNS and modified Fisher scales into ordered risk categories [6], and the SAHIT multinational models, which integrate clinical and radiological variables to predict functional outcome after aSAH [7]. More recently, machine learning approaches have been applied to large datasets to improve early DCI prediction; across these models, the variables most consistently retained include age, sex, clinical grade, hemorrhage burden, and treatment modality, though their incremental value over conventional regression methods and their external validity remain incompletely established [8]. Importantly, virtually this entire evidence base—both the individual risk factors and the prediction models derived from them—comes from populations living at or near sea level, and none of these tools account for the unique environmental and physiological exposures characteristic of chronically hypoxic high-altitude populations.

Populations residing permanently at high altitude differ from sea-level populations in several ways that could plausibly modify DCI pathophysiology. Chronic hypobaric hypoxia triggers compensatory erythrocytosis with progressive elevation in hemoglobin concentration, which increases blood viscosity, and is accompanied by remodeling of cerebrovascular reactivity and autoregulation. These physiological adaptations represent a double-edged sword: while they preserve systemic oxygen delivery, they may also reduce cerebrovascular reserve capacity [9,10]. Together with hypoxia-related endothelial dysfunction and hemostatic alterations, these changes have been implicated in the elevated burden of ischemic cerebrovascular disease observed at high altitude [10,11] and provide a strong biological rationale for hypothesizing that high-altitude residence might alter susceptibility to DCI following aneurysmal rupture [10]. Yet clinical evidence addressing this question remains remarkably sparse. The few existing studies of aSAH in high-altitude regions come almost entirely from the Tibetan Plateau and have focused primarily on surgical techniques and overall functional outcome rather than DCI as a primary endpoint [12,13]. Whether residential altitude, analyzed as a continuous exposure, is associated with DCI, and whether the compensatorily elevated hemoglobin levels of high-altitude dwellers influence DCI risk, have not been systematically investigated, leaving the interaction between altitude, hemoglobin, and secondary ischemic injury essentially unexplored.

Against this background, we conducted the present retrospective cohort study of aSAH patients managed at a single high-altitude referral center in Tibet with three prespecified objectives. First, we sought to describe the incidence and temporal distribution of DCI, adjudicated using the international consensus definition, in a cohort of permanent residents living at altitudes between approximately 2300 and 4900 m. Second, we examined the association between residential altitude—modeled as a continuous exposure to characterize the shape of any potential relationship—and admission hemoglobin with DCI occurrence. Third, we developed a parsimonious multivariable model to identify factors associated with DCI in this understudied population; notably, this model is intended as an etiological associational analysis rather than a bedside clinical prediction tool, as it includes post-treatment variables. Our analysis identified established markers of hemorrhage severity and treatment-related factors as the dominant associations, suggested a possible threshold-type increase in risk at the highest residential altitudes, and found no association between admission hemoglobin and DCI. These findings are intended to generate hypotheses and inform the design of future prospective surveillance studies of aSAH in high-altitude settings.

## 2. Methods

### 2.1 Study Design and Setting

We performed a single-center retrospective cohort study at the People’s Hospital of Tibet Autonomous Region (Lhasa, China), the primary tertiary referral center serving the Tibetan Plateau. Consecutive patients with aneurysmal subarachnoid hemorrhage admitted during [enrollment period] were screened for eligibility. We report the study in accordance with the Strengthening the Reporting of Observational Studies in Epidemiology (STROBE) statement [14]; a completed STROBE checklist is provided as Supplementary Material, and patient screening, exclusion, and analysis flow are depicted in Figure 1.

**Figure 1.**
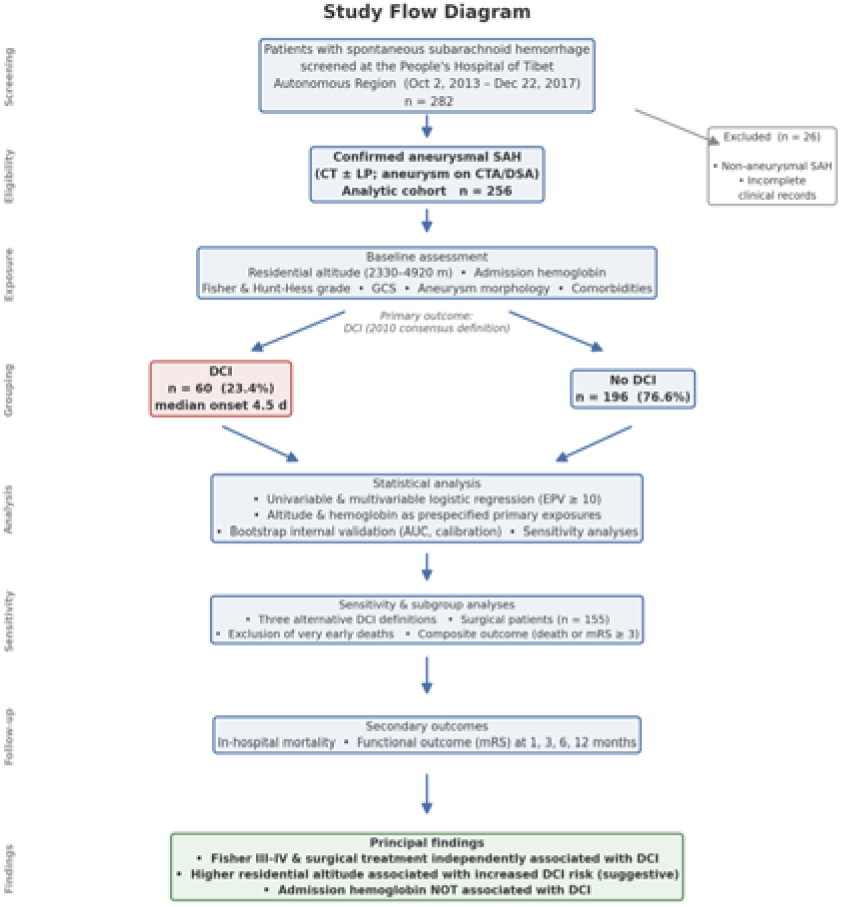
Patient enrollment flowchart (screening, exclusion, and analysis cohort). Placeholders ([total screened], [excluded]) are to be completed before submission.

The present analysis adopted the 2010 international consensus definition of DCI (Vergouwen et al. [3]) with adjudication of all events from medical records, and modeled residential altitude as a continuous exposure. The robustness of the altitude effect across alternative DCI definitions is addressed in the sensitivity analyses (Section 3.8) and in the Discussion.

### 2.2 Patient Population

Patients were eligible if they had a confirmed diagnosis of aSAH, established by non-contrast computed tomography (CT) (with or without lumbar puncture) demonstrating subarachnoid blood, together with CT angiography (CTA) or digital subtraction angiography (DSA) confirming at least one intracranial aneurysm. We excluded patients with non-aneurysmal subarachnoid hemorrhage (including perimesencephalic, traumatic, or vascular malformation–related bleeding) and those with incomplete clinical records. After applying these criteria, 256 patients formed the final analytic cohort. All patients resided on the Tibetan Plateau at residential altitudes ranging from 2330 to 4920 m. As the center functions as the regional referral hub, patients who died before transfer or within hours of arrival before diagnostic workup are underrepresented; accordingly, the cohort is best characterized as a referred, predominantly good-grade, surgical-capable population (see Limitations).

### 2.3 Definition of DCI and Adjudication

The primary outcome was delayed cerebral ischemia (DCI), defined according to the 2010 international consensus definition proposed by Vergouwen et al. [3] as new-onset cerebral ischemia occurring during hospitalization that was not attributable to other causes. Operationally, DCI required (i) clinical deterioration—defined as a new focal neurological deficit or a decrease of at least 2 points on the Glasgow Coma Scale (GCS)—persisting for at least 1 hour, not apparent immediately after aneurysm occlusion, and not attributable to other causes (e.g., procedural infarction related to aneurysm treatment, hypoxic-ischemic injury at presentation, hydrocephalus, rebleeding, seizures, electrolyte disturbance, or sedation), and/or (ii) a new ischemic lesion on follow-up CT not present on the admission or early post-procedural scan and not attributable to other causes. DCI events were adjudicated retrospectively from the medical records. For each event, the adjudication basis was categorized as symptoms plus CT, symptoms only, or CT only, and the time of onset was recorded in days from ictus. Events were adjudicated independently by two investigators, with disagreements resolved by consensus; because the residential address appears in the medical record, adjudicators could not be fully blinded to altitude, and formal inter-rater reliability (κ) was not quantified—both are acknowledged as limitations. Follow-up CT was obtained on clinical indication rather than under a fixed surveillance protocol, so CT-only events are likely under-ascertained.

### 2.4 Variable Collection

We abstracted demographic characteristics (age, sex), vascular risk factors (hypertension, smoking, alcohol consumption), admission clinical severity (GCS, Hunt-Hess grade, Fisher grade), aneurysm characteristics (maximum diameter, aspect ratio, size ratio, irregularity index, posterior circulation location, multiplicity, and giant aneurysm defined as ≥25 mm), treatment variables (surgical treatment, interval from symptom onset to aneurysm securing [onset-to-surgery interval], tirofiban use, and antiplatelet agents), and admission hemoglobin from medical records. For regression analyses, admission severity scores were additionally dichotomized (Hunt-Hess grade III–IV, Fisher grade III–IV, and GCS ≤ 8).

Surgical treatment was defined as definitive aneurysm occlusion by either microsurgical clipping or endovascular coiling; patients managed conservatively (without aneurysm occlusion) were classified as non-surgical. The specific occlusion technique (clipping versus coiling) was not uniformly recorded in a manner amenable to analysis and therefore could not be modeled separately; this represents an unmeasured source of confounding, as clipping has been associated with higher DCI risk than coiling [15]. The high rate of tirofiban and antiplatelet use, confined exclusively to surgically treated patients, suggests that a substantial proportion underwent endovascular treatment.

Residential altitude, our primary exposure of interest, was determined by matching each patient’s registered residential address to a geographic database and extracting the corresponding elevation. For the 18 patients born in mainland China who had resided in Tibet long-term, we used the altitude of their actual long-term residence rather than their birthplace, as physiological acclimatization and chronic hypoxic exposure reflect the true residential environment. Registered residential address remains an imperfect surrogate for chronic hypoxic dose (see Limitations). During data cleaning, we identified one aneurysm with an implausible recorded size (260/120 mm) and corrected it to 26.0/12.0 mm based on its maximum diameter; no other extreme morphometric values required correction on review.

### 2.5 Outcomes

The primary outcome was DCI. Secondary outcomes included in-hospital mortality and functional outcome, assessed using the modified Rankin Scale (mRS) [16] at 1, 3, 6, and 12 months. Poor functional outcome was defined as an mRS score of 3 or higher. Patients who died were excluded from mRS denominators at each time point (i.e., deaths were not coded as mRS 6); consequently, mRS-based proportions reflect survivors only. To address this, we additionally analyzed a composite outcome of death or mRS ≥3 (with death counted as the worst possible state) as a sensitivity measure.

### 2.6 Statistical Analysis

We assessed normality of continuous variables using the Shapiro-Wilk test. Normally distributed variables are presented as mean ± standard deviation (SD) and compared using the independent-samples t-test; non-normally distributed variables are presented as median (interquartile range [IQR]) and compared using the Mann-Whitney U test. Categorical variables are presented as n (%) and compared using the chi-square test (with continuity correction) or Fisher’s exact test when any expected cell count was below 5.

Univariable logistic regression was used to screen candidate predictors of DCI, and odds ratios (ORs) with 95% confidence intervals (CIs) were estimated. Residential altitude and admission hemoglobin were designated a priori as the primary exposures and were forced into every multivariable model regardless of their univariable significance; all remaining covariates were selected on the basis of established clinical relevance and the prior literature rather than univariable p-values. Clinically relevant variables were entered into multivariable logistic regression models, and adjusted ORs (aORs) with 95% CIs were reported. To avoid overfitting, the number of covariates was restricted to maintain an events-per-variable (EPV) ratio of at least 10, in accordance with the rule proposed by Peduzzi et al. [17].

We assessed multicollinearity using variance inflation factors (VIFs). Because Hunt-Hess grade, Fisher grade, and GCS were mutually collinear (VIF 2.07–4.32), we did not enter Hunt-Hess and Fisher grades into the same model and instead constructed alternative models. The prespecified primary (confirmatory) model was Model A, which included Fisher grade, surgical treatment, altitude, and hemoglobin (Model A covariate VIFs were low, ranging from 1.01–1.04). Model B substituted Hunt-Hess grade for Fisher grade and added multiple aneurysms. Model C examined the effect of onset-to-surgery interval and was therefore restricted to surgically treated patients (n = 155), as onset-to-surgery interval is a time-to-treatment exposure defined only for patients who underwent aneurysm securing; non-surgical patients have no such exposure, and assigning them an interval value would conflate the decision to operate with operative timing. Models A (primary) and B were fitted to the full cohort. This structure prespecifies Model A as the primary model, with Models B and C serving as secondary/sensitivity specifications.

Model performance was quantified using the area under the receiver-operating-characteristic curve (AUC), with 95% CIs derived from 2000 bootstrap resamples, and calibration (Brier score and calibration slope). We performed several sensitivity analyses: (i) robustness of the altitude effect across alternative DCI definitions; (ii) a formal test for linear trend in DCI incidence across ordered altitude strata (altitude modeled as a linear term) to evaluate the linear dose–response assumption; and (iii) a competing-risk sensitivity analysis excluding the two patients who died very early (within 3 days of admission, before the DCI window opened) to assess the influence of early death as a competing event on the altitude estimate. Because complete death-timing data were unavailable, we could not fit a formal Fine–Gray subdistribution hazard model, which we acknowledge as a limitation. We performed no imputation for missing data; analyses were based on complete cases, with denominators reported for each analysis (onset-to-surgery interval was unavailable for one surgical patient, yielding n = 155 in Model C, and numbers of patients with available assessments are shown for each follow-up time point in Table 4). We applied no adjustment for multiple comparisons; all p values are nominal, and analyses should be regarded as exploratory/hypothesis-generating. All tests were two-sided, with statistical significance set at α = 0.05. Analyses were performed in Python 3.12 using the pandas, scipy, statsmodels, and scikit-learn packages.

### 2.7 Sensitivity Analyses

The robustness of the altitude effect across alternative DCI definitions was examined as a prespecified sensitivity analysis. The present analysis adopted the 2010 international consensus definition of DCI (Vergouwen et al. [3]), with events adjudicated from medical records as described in Section 2.3. Three definitions were compared: (1) the primary consensus definition (in-hospital cerebral ischemia); (2) a more stringent definition requiring both clinical symptoms and CT confirmation; and (3) the stringent definition further restricted to events within the canonical 4–14-day window. The altitude point estimate was stable across definitions (adjusted OR 1.08–1.10 per 100 m); however, under the strictest definition the number of events was limited (39 events, EPV 9.8) and the confidence interval crossed 1.0, indicating that this stricter analysis is underpowered and that the corresponding findings should be interpreted with caution. Point-estimate stability across definitions therefore reflects a consistent direction of association but does not by itself establish adequate statistical power.

### 2.8 Ethics and Reporting

The study protocol was approved by the Ethics Committee of the People’s Hospital of Tibet Autonomous Region (XZ2024115). The requirement for individual informed consent was waived due to the retrospective design and use of de-identified data. Reporting adheres to the STROBE guideline [14].

### 2.9 Data Availability

The de-identified analytic dataset and analysis code are not publicly available due to patient privacy restrictions but are available from the corresponding author upon reasonable request and with appropriate institutional approval.

### 2.10 Funding and Conflicts of Interest

This research was supported by the Natural Science Foundation of the Tibet Autonomous Region,

Project Number: XZ2024ZR-ZY004 (Z). There is no conflict of interest among the authors of this article.

## 3. Results

### 3.1 Cohort Characteristics

A total of 256 aSAH patients met eligibility criteria and were included in the analysis (Figure 1); all resided at altitudes between 2330 and 4920 m. DCI developed in 60 patients (23.4%). Median time from ictus to DCI onset was 4.5 days (IQR 4.0–6.8; range 1–18 days). Among the 58 patients with recorded onset times, 43 (74.1%) developed DCI within the 4–14 day window; the remaining 15 events (25.9%) occurred outside this window, a deviation from the canonical DCI time course that we examine in sensitivity analyses (Section 3.8) and discuss as a limitation. Two events lacked recorded onset times. The adjudication basis was symptoms plus CT in 52 patients, symptoms only in 6, and CT only in 2. The observed DCI incidence was somewhat lower than the approximately 30% rate reported in contemporary aSAH guidelines and population series [1], a difference that likely reflects retrospective case ascertainment, the predominance of good-grade referred patients, and early deaths occurring before the DCI window (competing risk), rather than choice of definition alone.

### 3.2 Baseline Characteristics

Baseline characteristics stratified by DCI status are presented in Table 1. Compared with the non-DCI group, patients who developed DCI had significantly lower admission GCS, higher Hunt-Hess and Fisher grades, a higher frequency of GCS ≤ 8, and a higher rate of surgical treatment (all p < 0.001); across the full cohort (with non-surgical patients assigned an interval of 0), onset-to-surgery interval was also longer in the DCI group (p < 0.001), though this largely reflects the surgical/non-surgical contrast rather than surgical timing alone (see Table 1 footnote). In contrast, age, sex, admission hemoglobin, and residential altitude did not differ significantly between groups on unadjusted comparison. Variables marked with † (in-hospital death, total hospital stay, ICU days, nimodipine days, and mannitol days) represent downstream consequences of DCI rather than antecedent predictors and were excluded from risk-factor modeling.

**Table 1.**
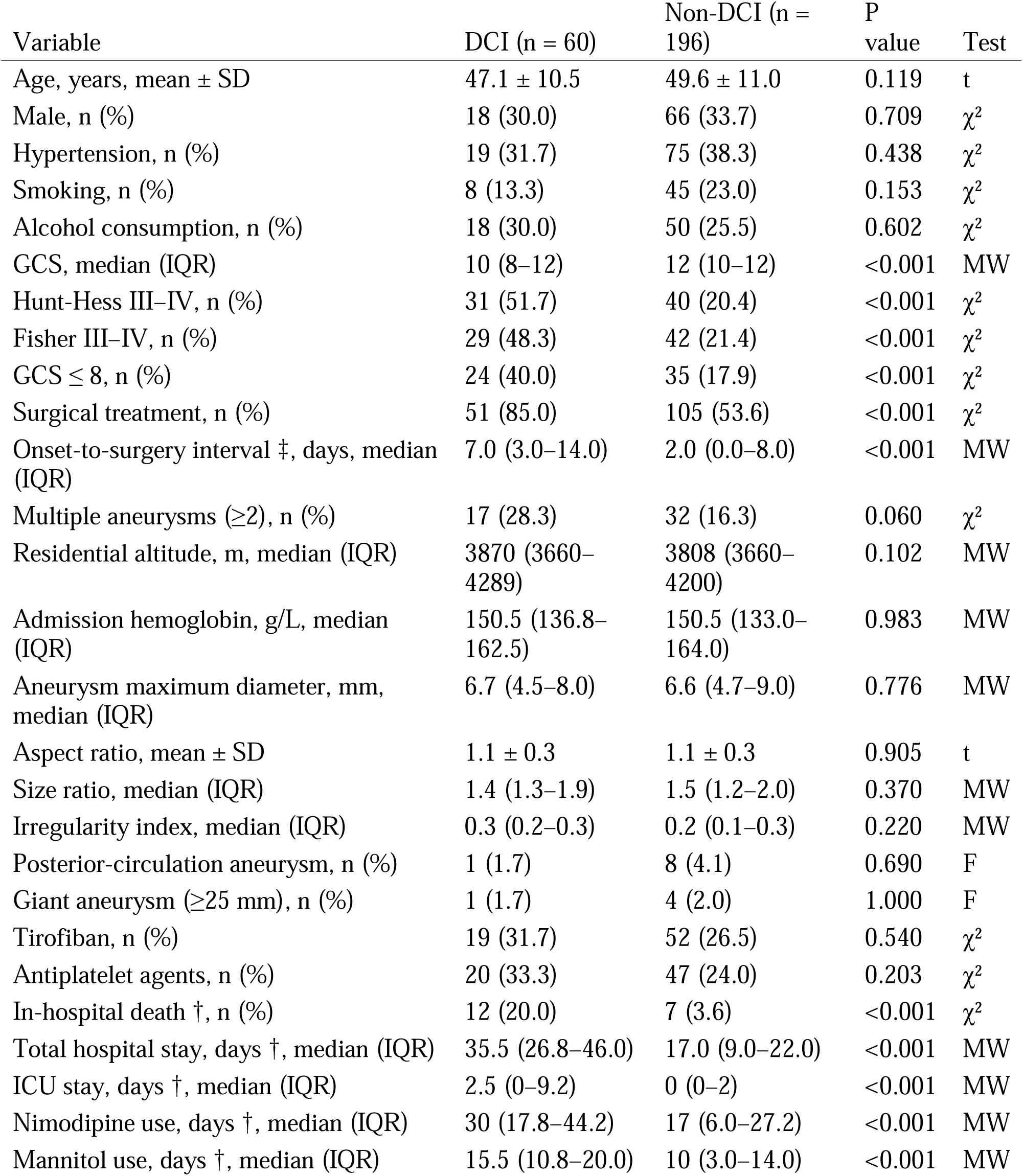
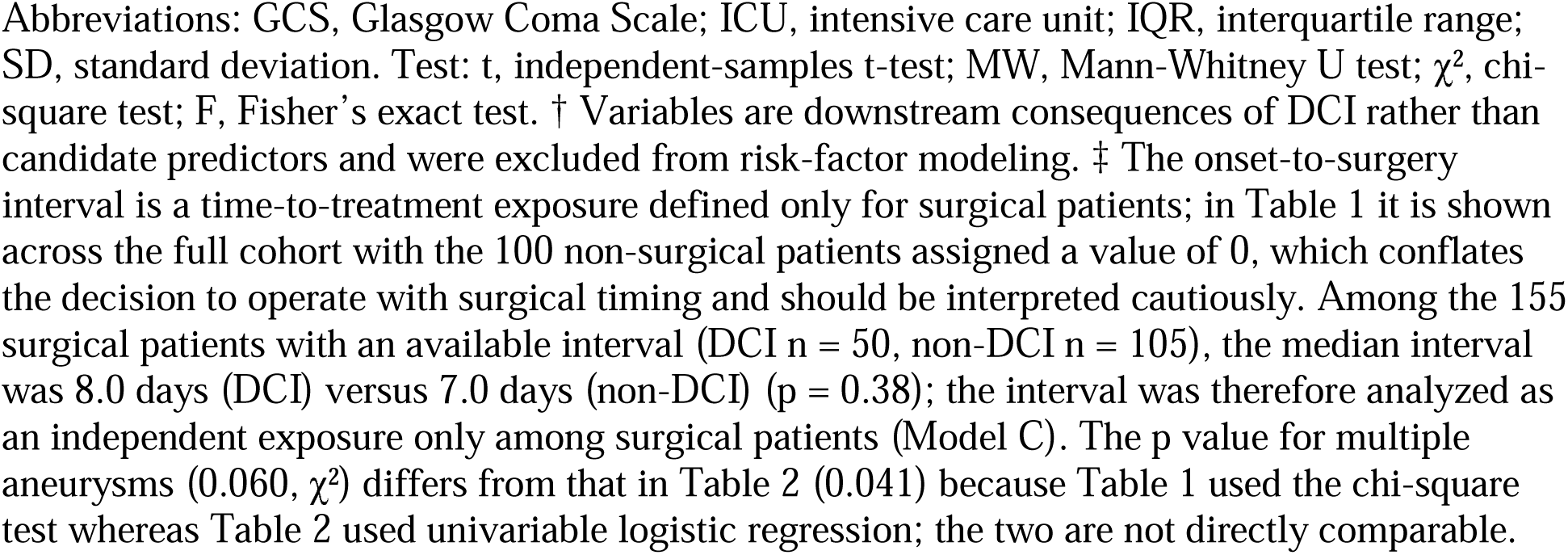
Baseline characteristics of patients with and without delayed cerebral ischemia (DCI).

Baseline comparisons revealed a consistent pattern of greater initial clinical and radiological severity among patients who subsequently developed DCI. Admission GCS was significantly lower in the DCI group (median 10 vs 12, p < 0.001), and both Hunt-Hess and Fisher scales showed a marked shift toward higher grades, with high-grade (III–IV) scores more than twice as frequent in the DCI group (51.7% vs 20.4% and 48.3% vs 21.4%, respectively). Surgical treatment was substantially more common among patients with DCI (85.0% vs 53.6%). Across the full cohort (non-surgical patients assigned an interval of 0), onset-to-surgery interval was longer in the DCI group (median 7.0 vs 2.0 days); however, among surgical patients themselves the interval was similar between groups (median 8.0 vs 7.0 days, p = 0.38), indicating the univariable interval contrast largely reflects the surgical/non-surgical difference rather than surgical timing per se—an observation that motivated restricting interval analysis to surgical patients in Model C. Notably, age, sex, admission hemoglobin, and residential altitude were balanced between groups on unadjusted comparison, as were aneurysm size and morphometric indices. The † variables represent consequences rather than causes of DCI and were therefore not considered candidate risk factors.

### 3.3 Univariable Analysis

Results of univariable logistic regression are presented in Table 2. Six variables were significantly associated with DCI: Hunt-Hess grade III–IV, Fisher grade III–IV, GCS (per point), GCS ≤ 8, surgical treatment, and multiple aneurysms. Residential altitude showed a positive but non-significant trend, while admission hemoglobin was not associated with DCI.

**Table 2.** Univariable logistic regression for delayed cerebral ischemia (n = 256; 60 events).

| Variable | OR (95% CI) | P value |
| --- | --- | --- |
| Age (per 1 year) | 0.98 (0.95–1.01) | 0.126 |
| Male | 0.84 (0.45–1.58) | 0.596 |
| Hypertension | 0.75 (0.40–1.38) | 0.354 |
| Smoking | 0.52 (0.23–1.17) | 0.112 |
| Alcohol consumption | 1.25 (0.66–2.37) | 0.491 |
| Hunt-Hess III–IV | 4.17 (2.26–7.70) | <0.001 |
| Fisher III–IV | 3.43 (1.86–6.32) | <0.001 |
| GCS (per 1 point) | 0.76 (0.68–0.85) | <0.001 |
| GCS ≤ 8 | 3.07 (1.63–5.77) | <0.001 |
| Surgical treatment | 4.91 (2.29–10.52) | <0.001 |
| Multiple aneurysms (≥2) | 2.03 (1.03–3.99) | 0.041 |
| Aneurysm maximum diameter (per 1 mm) | 1.00 (0.94–1.05) | 0.892 |
| Irregularity index (per 0.1) | 1.08 (0.93–1.26) | 0.319 |
| Size ratio (per 0.5) | 0.86 (0.68–1.10) | 0.228 |
| Admission hemoglobin (per 10 g/L) | 1.00 (0.91–1.11) | 0.939 |
| Residential altitude (per 100 m) | 1.06 (0.98–1.13) | 0.134 |
| Posterior-circulation aneurysm | 0.40 (0.05–3.25) | 0.390 |
| Tirofiban | 1.28 (0.68–2.41) | 0.437 |
| Antiplatelet agents | 1.59 (0.85–2.97) | 0.151 |

Abbreviations: CI, confidence interval; GCS, Glasgow Coma Scale; OR, odds ratio. The odds ratio for posterior-circulation aneurysm is based on only 9 patients and is therefore imprecise. All p values are nominal (no adjustment for multiple comparisons).

Univariable logistic regression identified six variables significantly associated with DCI, all reflecting either baseline neurological severity or treatment-related factors. The strongest unadjusted associations were observed for surgical treatment (OR 4.91), Hunt-Hess III–IV (OR 4.17), Fisher III–IV (OR 3.43), and GCS ≤ 8 (OR 3.07), while each one-point increase in GCS was protective (OR 0.76). Multiple aneurysms conferred an approximately twofold increase in DCI odds (OR 2.03, p = 0.041). Our primary exposure of interest, residential altitude, showed a positive but non-significant association (OR 1.06 per 100 m, p = 0.134). In contrast, admission hemoglobin was entirely neutral (OR 1.00 per 10 g/L, p = 0.939), providing no support for a hemoglobin-mediated mechanism at the univariable level. Demographic characteristics, comorbidities, aneurysm size and morphology, aneurysm location, and use of tirofiban or antiplatelet agents were not significantly associated with DCI.

### 3.4 Multivariable Analysis

Multivariable models are summarized in Table 3 and visualized in Figure 2. In the prespecified primary Model A (n = 256; EPV 15.0; AUC 0.759 [bootstrap 95% CI 0.690–0.828]), Fisher grade III–IV and surgical treatment were independently associated with DCI, and the altitude association was positive but fell just short of conventional significance (p = 0.051). In the alternative Model B (n = 256; AUC 0.710), Hunt-Hess III–IV and multiple aneurysms were significant, while altitude was attenuated and non-significant (p = 0.157). In Model C, restricted to surgical patients (n = 155; 50 events; EPV 12.5; AUC 0.731) and incorporating onset-to-surgery interval, both interval and residential altitude were independently associated with DCI (altitude p = 0.021). Across these specifications the altitude point estimate remained stable (∼1.06–1.11 per 100 m) but its statistical significance did not (p ranging from 0.021 to 0.157), and admission hemoglobin was not independently associated with DCI in any model.

**Figure 2.**
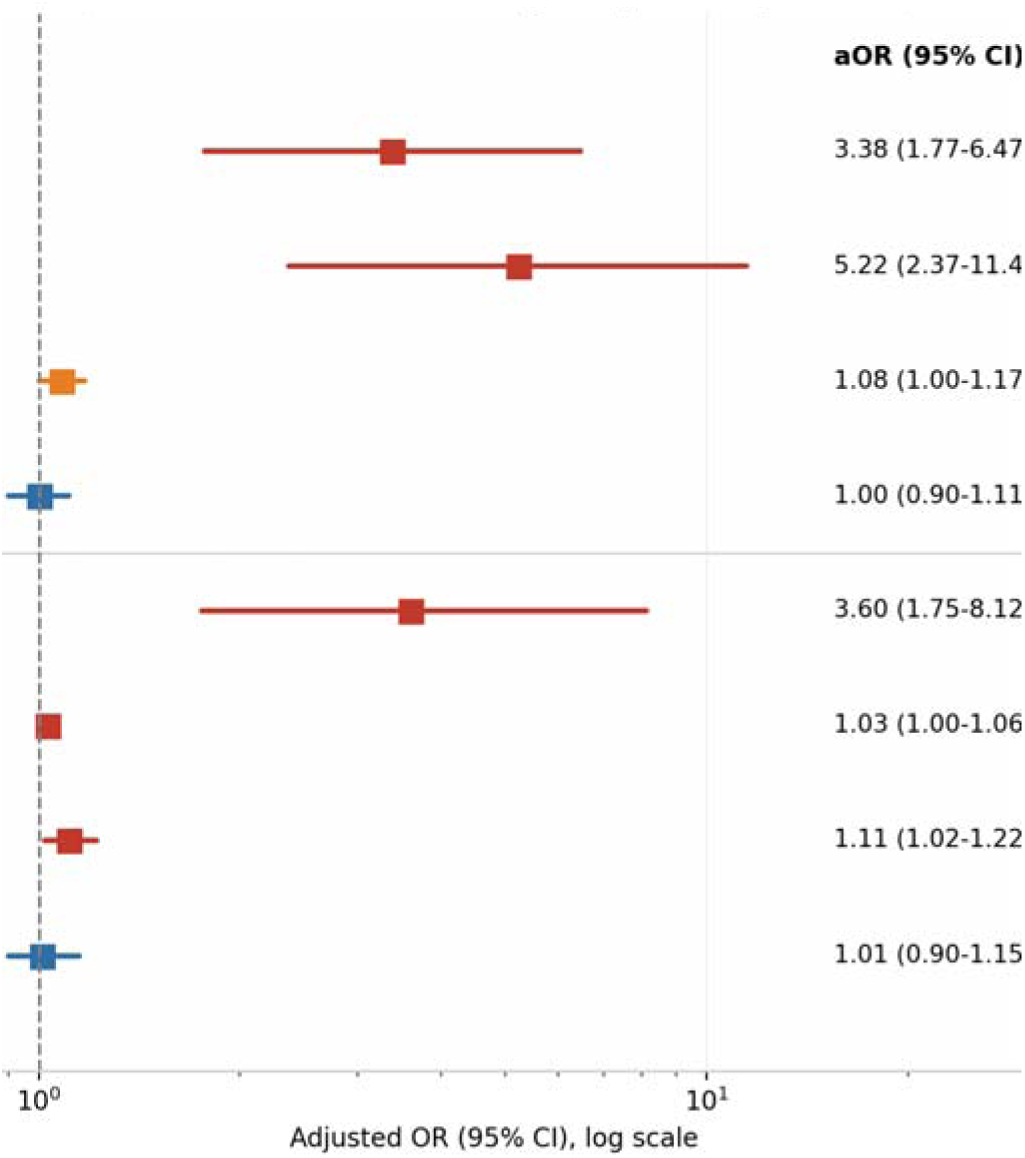
Forest plot of adjusted odds ratios (aOR, 95% CI) from the prespecified primary Model A (full cohort) and Model C (restricted to surgical patients, n = 155). Fisher grade and surgical treatment were significantly associated with DCI; the altitude association was borderline in Model A (p = 0.051) and significant among surgical patients (Model C, p = 0.021); hemoglobin was null in both.

**Table 3.** Multivariable logistic regression models for delayed cerebral ischemia.

| Model | Variable | aOR (95% CI) | P value |
| --- | --- | --- | --- |
| **Model A** (primary; full cohort, n = 256; EPV 15.0; AUC 0.759 [0.690–0.828]) | Fisher III–IV | 3.38 (1.77–6.47) | <0.001 |
|  | Surgical treatment | 5.22 (2.37–11.47) | <0.001 |
|  | Residential altitude (per 100 m) | 1.08 (1.00–1.17) | 0.051 |
|  | Admission hemoglobin (per 10 g/L) | 1.00 (0.90–1.11) | 0.988 |
| **Model B** (full cohort, n = 256; EPV 15.0; AUC 0.710) | Hunt-Hess III–IV | 4.45 (2.37–8.36) | <0.001 |
| | Multiple aneurysms ($\geq 2$) | 2.12 (1.03–4.38) | 0.042 |
|  | Residential altitude (per 100 m) | 1.06 (0.98–1.14) | 0.157 |
|  | Admission hemoglobin (per 10 g/L) | 1.00 (0.90–1.11) | 0.960 |
| **Model C** (surgical patients only, n = 155; 50 events; EPV 12.5; AUC 0.731) | Fisher III–IV | 3.60 (1.75–8.12) | 0.001 |
|  | Onset-to-surgery interval (per 1 day) | 1.030 (1.004–1.058) | 0.026 |
|  | Residential altitude (per 100 m) | 1.11 (1.02–1.22) | 0.021 |
|  | Admission hemoglobin (per 10 g/L) | 1.01 (0.90–1.15) | 0.75 |

*Abbreviations: aOR, adjusted odds ratio; AUC, area under the receiver-operating-characteristic curve; CI, confidence interval; EPV, events per variable. Model A is the prespecified primary model; Models B and C are secondary/sensitivity specifications. Models A and B were fitted to the full cohort; residential altitude and hemoglobin were forced into all models as prespecified primary exposures. Model C was restricted to surgically treated patients because the onset-to-surgery interval is a time-to-treatment exposure defined only for patients who underwent aneurysm securing; non-surgical patients have no such exposure. Model A covariate VIFs ranged from 1.01 to 1.04; Hunt-Hess grade, Fisher grade, and GCS were mutually collinear (VIF 2.07–4.32), which is why they were not entered into the same model. AUC 95% CI is shown for the primary Model A (2000 bootstrap resamples).*

Multivariable modeling confirmed baseline radiological severity and surgical treatment as the dominant independent associations while clarifying the role of residential altitude. In primary Model A, Fisher grade III–IV (aOR 3.38) and surgical treatment (aOR 5.22) remained strongly and independently associated with DCI, with adequate discrimination (AUC 0.759) and negligible multicollinearity (VIF 1.01–1.04). The altitude effect was positive but fell just short of conventional significance (aOR 1.08 per 100 m, p = 0.051). In Model B, which substituted Hunt-Hess grade for Fisher grade, Hunt-Hess III–IV (aOR 4.45) and multiple aneurysms (aOR 2.12) were significant, while altitude was attenuated (aOR 1.06, p = 0.157) and hemoglobin remained non-significant (aOR 1.00, p = 0.960). In Model C, restricted to surgical patients and adjusted for onset-to-surgery interval, each additional day to surgery was associated with higher DCI odds (aOR 1.030, p = 0.026), and the altitude association was stronger and statistically significant (aOR 1.11 per 100 m, p = 0.021); however, because Model C conditions on having undergone surgery—a post-baseline, potentially collider-affected state—this estimate should be interpreted as an association among surgical patients rather than a cohort-level effect. Admission hemoglobin was not independently associated with DCI in any model.

### 3.5 Altitude-Stratified Incidence

DCI incidence across successive altitude strata was: <3600 m, 5/25 (20.0%); 3600–4000 m, 30/140 (21.4%); 4000–4400 m, 13/44 (29.5%); and >4400 m, 12/47 (25.5%) (Figure 3). The distribution was non-monotonic: rates rose to a peak in the 4000–4400 m stratum and then declined slightly in the highest (>4400 m) stratum. A formal test for linear trend across ordered strata (altitude modeled as a linear term) was non-significant (p = 0.385), indicating the data do not support a strictly linear dose–response relationship. When strata above 4000 m were combined, DCI incidence was 25/91 (27.5%), compared with 35/165 (21.2%) at altitudes of 4000 m or below (chi-square p = 0.328); although numerically higher above 4000 m, this difference did not reach statistical significance. These patterns are consistent with a possible threshold-type increase in risk at the highest altitudes, but small stratum sizes (25–47 patients) render stratum-specific estimates imprecise, and the shape of the altitude–DCI relationship should be regarded as exploratory.

**Figure 3.**
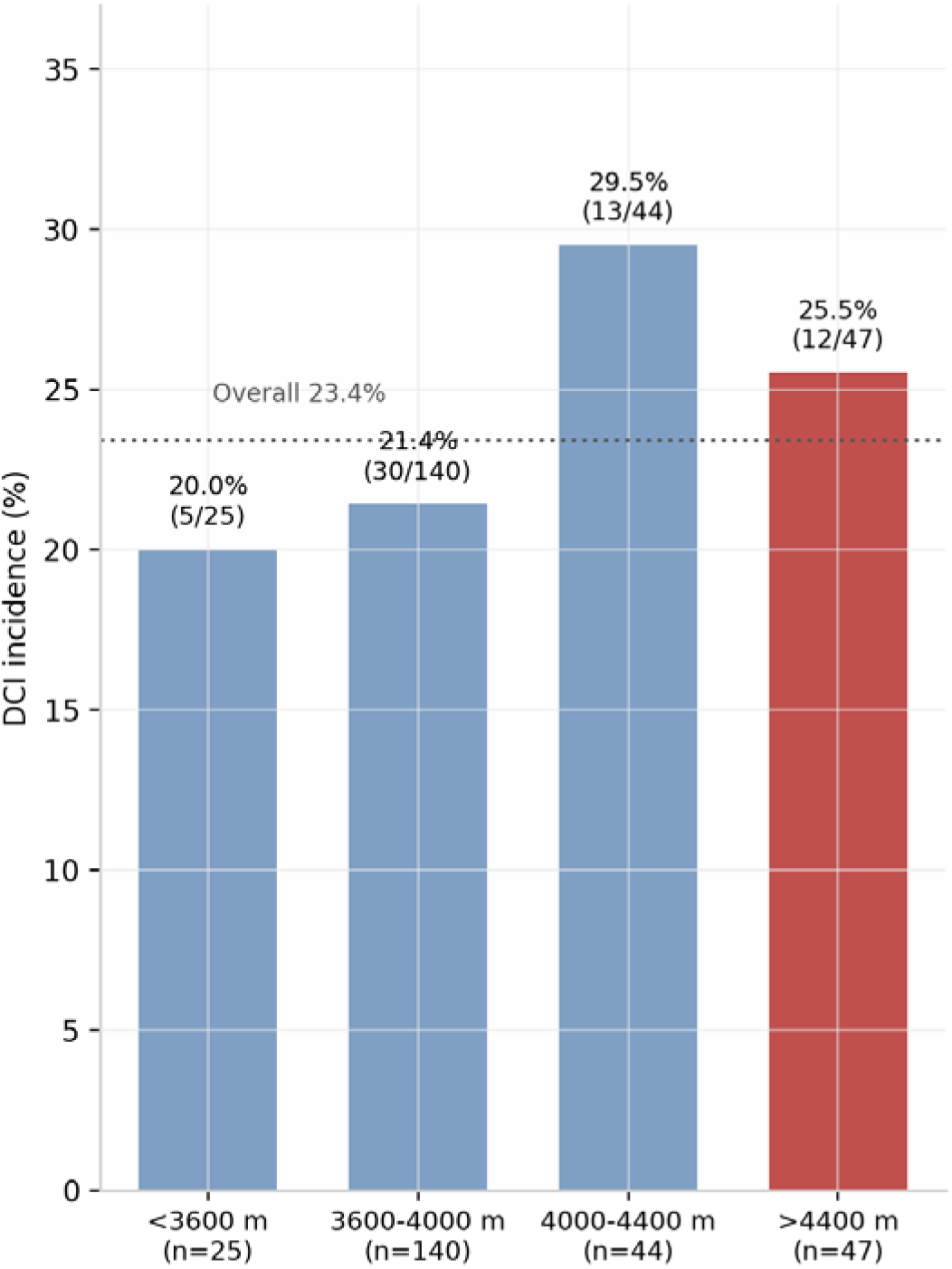
DCI incidence by altitude stratum. The distribution was non-monotonic (20.0% → 21.4% → 29.5% → 25.5%), and a formal test for linear trend was non-significant (p = 0.385), arguing against a strictly linear dose–response.

### 3.6 DCI and Clinical Outcomes

The association between DCI and clinical outcomes is shown in Table 4. DCI was associated with markedly worse in-hospital and long-term outcomes. In-hospital mortality was significantly higher in the DCI group, and poor functional outcome (mRS ≥ 3) was significantly more frequent at every follow-up point among survivors. Because patients who died were excluded from mRS denominators, we additionally analyzed a composite outcome of death or mRS ≥ 3 at 1 year: this was present in 35/60 (58.3%) of the DCI group versus 35/196 (17.9%) of the non-DCI group (OR 6.33, 95% CI 3.39–11.83).

**Table 4.**
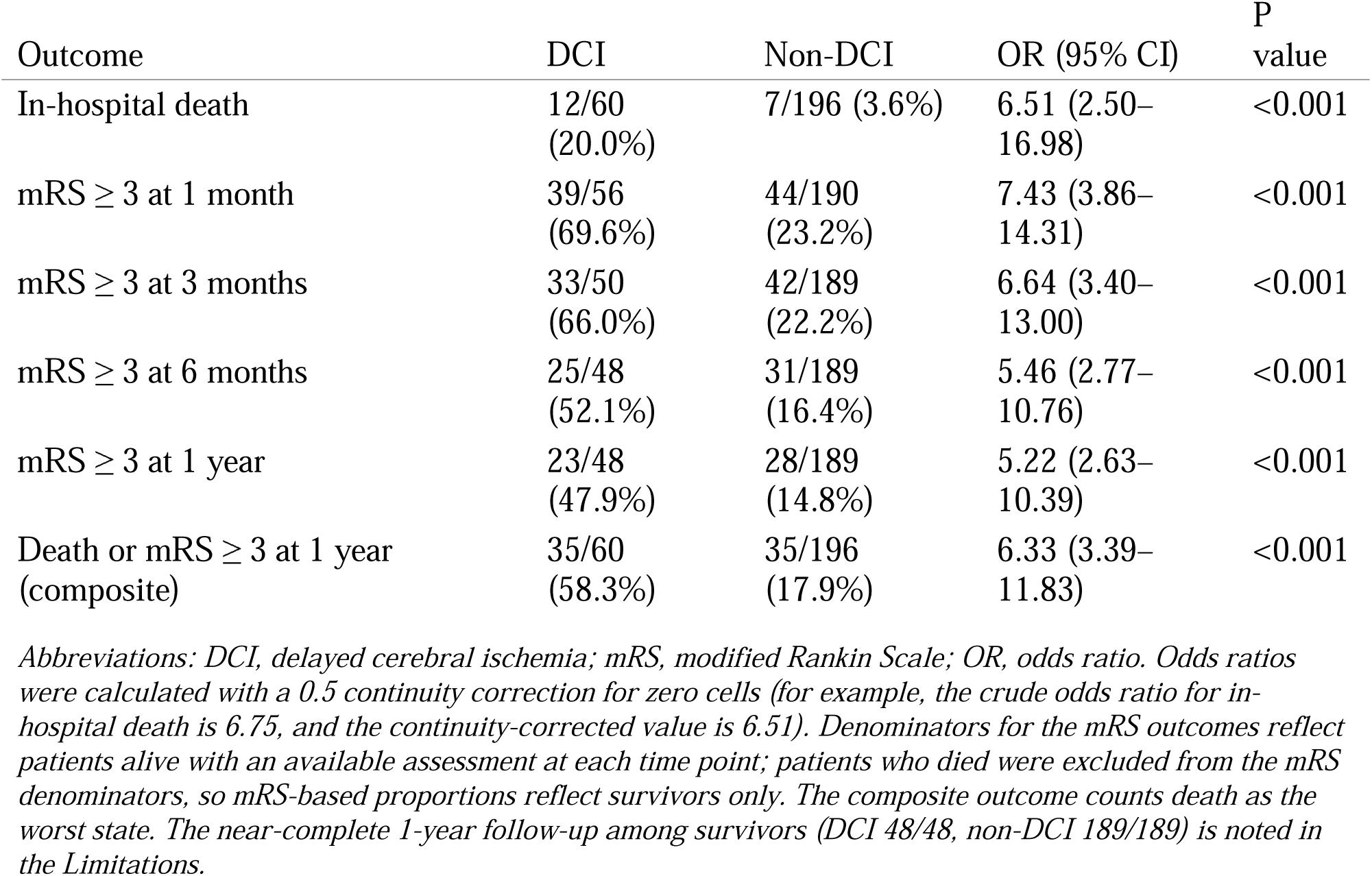
Association between delayed cerebral ischemia and clinical outcomes.

DCI was strongly and consistently associated with adverse outcomes across the entire follow-up period. In-hospital mortality was nearly sixfold higher among patients with DCI (20.0% vs 3.6%; OR 6.51). The excess in poor functional outcome (mRS ≥ 3) among survivors was even more pronounced and persisted over time: odds of an unfavorable outcome were approximately sevenfold higher at 1 month (OR 7.43) and remained more than fivefold higher at 3 months (OR 6.64), 6 months (OR 5.46), and 1 year (OR 5.22). Although the absolute proportion of surviving patients with mRS ≥ 3 declined over time in both groups—consistent with partial neurological recovery—the relative disadvantage conferred by DCI did not attenuate, and at 1 year almost half of the surviving DCI group (47.9%) remained functionally dependent, compared with only 14.8% of the non-DCI group. When death was incorporated as the worst state (composite of death or mRS ≥ 3), 58.3% of the DCI group had an unfavorable 1-year outcome versus 17.9% of the non-DCI group (OR 6.33). All these associations were highly significant (p < 0.001), underscoring the strong and consistent relationship between DCI and both early mortality and long-term disability in this high-altitude aSAH cohort.

### 3.7 Model Performance, Calibration, and Validation

Model performance is shown in Figure 4. The prespecified primary Model A achieved acceptable discrimination (AUC 0.759, bootstrap 95% CI 0.690–0.828). We assessed calibration using the Brier score (0.152) and calibration slope (1.13), the latter indicating mild overfitting/shrinkage consistent with the modest events-per-variable ratio; the calibration curve tracked the identity line reasonably closely across the observed risk range (Figure 4b). Because the model incorporates post-treatment variables (surgical treatment and, in Model C, onset-to-surgery interval), it represents an etiological/associational model and is not intended or validated as a clinical prediction model for use at admission; its coefficients quantify associations rather than providing a deployable risk score. If admission-time risk stratification were the objective, a prediction model would need to exclude post-treatment variables, be reported in accordance with TRIPOD guidelines, and undergo external validation—none of which was the aim of the present analysis.

**Figure 4.**
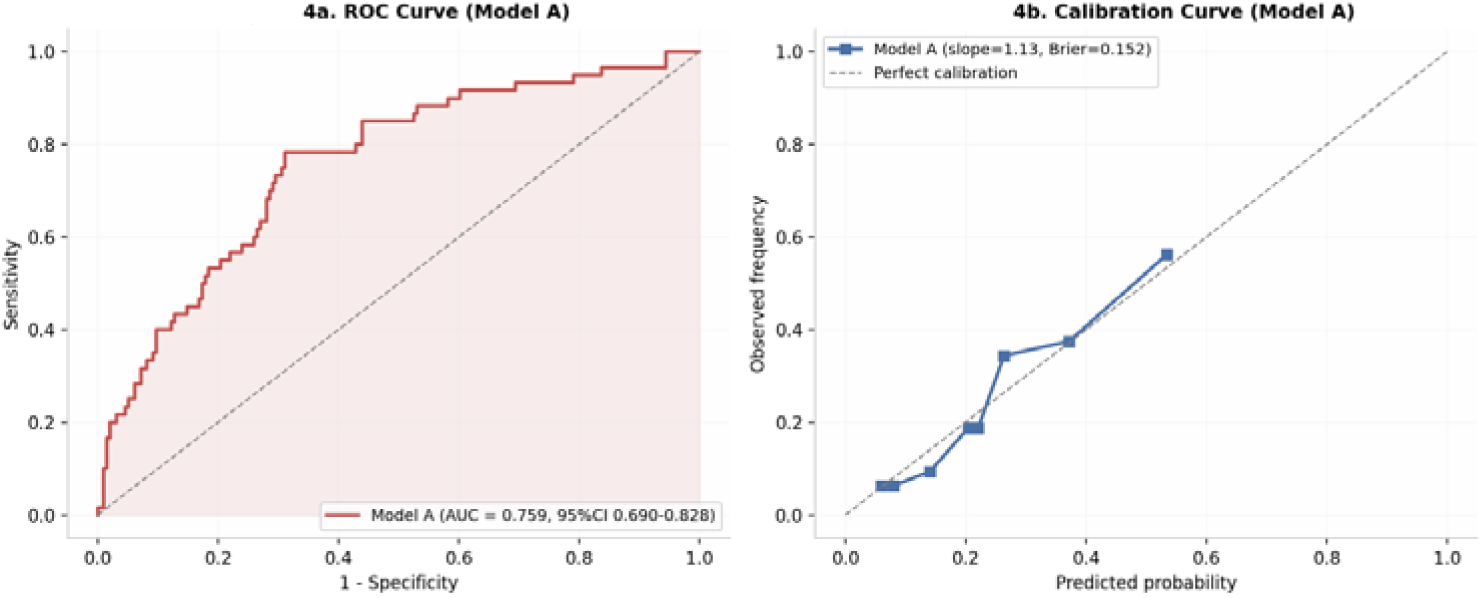
(a) Receiver-operating-characteristic curve for the primary Model A (AUC 0.759, bootstrap 95% CI 0.690–0.828). (b) Calibration curve for Model A (calibration slope 1.13, Brier score 0.152); the dashed line indicates perfect calibration.

### 3.8 Sensitivity Analyses

The robustness of the altitude effect across alternative DCI definitions is shown in Supplementary Table S1. The altitude point estimate remained stable across definitions. Using the primary definition (60 events, 23.4%; EPV 15.0), the adjusted OR for altitude was 1.08 per 100 m (p = 0.051). Using the more stringent definition requiring both clinical symptoms and CT confirmation (52 events, 20.3%; EPV 13.0), the adjusted OR was 1.10 per 100 m (p = 0.025). Restricting events to the 4–14 day window (39 events, 15.2%; EPV 9.8) yielded an adjusted OR of 1.08 per 100 m (p = 0.075), though this strictest analysis was underpowered (EPV < 10, confidence interval crossing 1.0). In a competing-risk sensitivity analysis excluding the two patients who died within 3 days of admission (before the DCI window opened), the altitude estimate was essentially unchanged (aOR 1.08 per 100 m, 95% CI 1.00–1.17; p = 0.050), indicating the borderline altitude association is not materially driven by very early deaths. Across consensus-based definitions, Fisher grade III–IV and surgical treatment remained significantly associated with DCI, and admission hemoglobin was not significant (p > 0.4). The consistency o the altitude point estimate (adjusted OR 1.08–1.10 per 100 m) indicates a stable direction of association, but fluctuating significance and limited power under stricter definitions mean the altitude finding should be considered suggestive rather than definitive.

**Supplementary Table S1.** Sensitivity analysis of the altitude effect across alternative definitions of delayed cerebral ischemia, and after exclusion of very early deaths.

| Analysis | Events | DCI rate | EPV | Altitude aOR per 100 m (95% CI) | P value |
| --- | --- | --- | --- | --- | --- |
| Primary: in-hospital cerebral ischemia (consensus) | 60 | 23.4% | 15.0 | 1.08 (1.00–1.17) | 0.051 |
| Symptoms plus CT (dual criteria) | 52 | 20.3% | 13.0 | 1.10 (1.01–1.19) | 0.025 |
| Symptoms plus CT, within 4–14 days | 39 | 15.2% | 9.8 | 1.08 (0.99–1.18) | 0.075 |
| Primary definition, excluding 2 deaths $\leq 3$ days (competing-risk sensitivity) | 60 | 23.6% | 15.0 | 1.08 (1.00–1.17) | 0.050 |

*Abbreviations: aOR, adjusted odds ratio; CI, confidence interval; CT, computed tomography; DCI, delayed cerebral ischemia; EPV, events per variable. All models include Fisher grade III–IV, surgical treatment, altitude, and hemoglobin (Model A specification). Across the consensus-based definitions, Fisher grade III–IV and surgical treatment remained significantly associated with DCI, and admission hemoglobin was not significant ( $p > 0.4$ ). The analysis restricted to the 4–14-day window is underpowered (EPV 9.8) and should be interpreted with caution.*

## 4. Discussion

In this single-center retrospective cohort of 256 patients with aneurysmal subarachnoid hemorrhage residing at altitudes of 2330–4920 m on the Tibetan Plateau, delayed cerebral ischemia developed in 60 patients (23.4%), with a median onset of 4.5 days and 74.1% of the 58 events with recorded onset times falling within the canonical 4–14-day window. Higher Fisher grade (III–IV) was independently associated with DCI (adjusted odds ratio [aOR] 3.38, 95% CI 1.77–6.47), as was surgical treatment (aOR 5.22). In the prespecified primary model, residential altitude was positively associated with DCI at approximately 8% per 100 m (aOR 1.08, 95% CI 1.00–1.17; p = 0.051), an estimate that remained numerically stable across the three consensus-based DCI definitions (aOR 1.08–1.10) but whose statistical significance fluctuated across model specifications (p = 0.021–0.157). Admission hemoglobin concentration, by contrast, showed no association with DCI in any model. DCI was strongly associated with both in-hospital mortality (OR 6.51) and poor 1-year functional outcome (composite of death or mRS ≥3: 58.3% vs 17.9%).

The robust association between Fisher grade and DCI aligns with the broader aSAH literature, in which subarachnoid blood volume and distribution rank among the most consistently replicated predictors of delayed ischemia [4,5,18]. Our adjusted estimate (aOR 3.38) falls within the range reported in systematic reviews and meta-analyses, and the effect was directionally identical—and somewhat larger—when Hunt–Hess grade III–IV was substituted for Fisher grade (aOR 4.45, 95% CI 2.37–8.36), indicating the signal reflects global disease severity rather than an idiosyncrasy of any single scale [4]. This interpretation is reinforced by the largest high-altitude aSAH cohort reported to date (n = 587), in which Hunt–Hess IV–V and modified Fisher III–IV independently predicted poor outcome with effect sizes (OR 3.33 and 3.19) closely mirroring our own [12]. The preservation of these canonical severity associations in a chronically hypoxic population suggests the cohort behaves in accordance with established aSAH pathophysiology, lending internal credibility to the more novel altitude-related findings that follow.

The most novel observation is the positive association between residential altitude and DCI. Altitude has not been incorporated as a candidate prognostic variable in established DCI prediction frameworks—neither the VASOGRADE scale nor the multinational SAHIT models considered environmental exposures, and a recent systematic review of machine learning models for early DCI prediction identified no altitude-related features [6,8]. To our knowledge, no prior study has reported an association between residential altitude modeled as a continuous exposure and DCI in this population. This claim requires qualification, however: the effect size was moderate, the association did not reach conventional significance in the prespecified primary model (p = 0.051) or in univariable analysis (p = 0.134), and statistical significance fluctuated across specifications (Model A p = 0.051; Model B p = 0.157; surgical-only Model C p = 0.021). We therefore regard the finding as suggestive rather than definitive.

Several convergent mechanisms lend biological plausibility to this association. Sustained hypobaric hypoxia activates hypoxia-inducible factor-1α, itself reported as an independent predictor of DCI [19]; impaired dynamic cerebral autoregulation strongly predicted DCI in a prospective sea-level study [20], and chronic hypobaric hypoxia is known to remodel cerebrovascular reactivity and autoregulation [9,10]; compensatory erythrocytosis increases blood viscosity, an established independent DCI predictor [21]; and hypoxia-responsive endothelin-1 signaling has been implicated in post-SAH vasospasm [22].

The shape of the altitude effect remains uncertain and, on present evidence, does not support a strictly linear dose–response relationship. Stratum-specific DCI rates were non-monotonic (20.0% below 3600 m, 21.4% at 3600–4000 m, 29.5% at 4000–4400 m, and 25.5% above 4400 m), the formal test for linear trend was non-significant (p = 0.385), and the difference between altitudes above versus at or below 4000 m (27.5% vs 21.2%) was not statistically significant (p = 0.328). A similarly weak altitude signal has been reported externally: in a large Andean cohort, residential altitude failed to predict cerebral arteriovenous malformation hemorrhage [23]. These patterns raise the hypothesis of a threshold-type relationship, in which risk accrues primarily above approximately 4000 m; however, we emphasize this is a post hoc, visually derived hypothesis that we did not formally fit or validate, stratum-specific estimates are imprecise (25–47 patients per stratum), and random fluctuation cannot be excluded. Whether the altitude–DCI relationship is genuinely threshold-type—a question best addressed with spline or change-point models in a larger cohort—represents a priority for future prospective studies rather than a conclusion of the present analysis.

Admission hemoglobin was unrelated to DCI in every model (all p > 0.85; median 150.5 g/L in both groups). This null finding is consistent with a meta-analysis of 40 studies including 14,701 patients concluding that hemoglobin at ictus carries no prognostic value and that only in-hospital nadirs are informative [24]. The high-hemoglobin end of the distribution represents an almost complete evidence gap, and to our knowledge no prior study has tested the hemoglobin–DCI hypothesis in a chronically altitude-adapted population. Our null result contrasts with a lowland registry study in which admission hemoglobin above 149.5 g/L was associated with higher in-hospital DCI (33.9% vs 22.0%) [25]. This discrepancy likely reflects the distinction between chronic compensatory and pathological erythrocytosis: thrombotic risk in polycythemia vera is driven by pan-myeloid activation rather than hemoglobin concentration per se [26], and hemoglobin above 152 g/L predicted in-hospital cerebral infarction in Tibetan patients with primary hemorrhagic neurovascular disease [27]. Hemoglobin dynamics may represent the more informative exposure—each 10 g/L (1 g/dL) in-hospital decrement was associated with DCI (OR 1.28) [28]—a dimension that a single admission measurement cannot capture. Finally, the transfusion literature requires careful interpretation. In anemic patients with aSAH, the SAHaRA trial found no functional outcome difference between liberal and restrictive transfusion strategies (33.5% vs 37.7% unfavorable neurological outcome; p = 0.22) [29]. In contrast, the TRAIN trial in acute brain injury (including an aSAH subgroup) found that a liberal strategy was associated with a lower rate of unfavorable neurological outcome at 180 days (62.6% vs 72.6%; adjusted RR 0.86, 95% CI 0.79–0.94; p = 0.002) and fewer cerebral ischemic events [30]. Critically, both trials enrolled anemic patients (hemoglobin thresholds ≤9–10 g/dL, i.e., ≤90–100 g/L), whereas our cohort had a median admission hemoglobin of 150.5 g/L and was, if anything, erythrocythemic. These randomized data therefore provide no direct evidence on hemoglobin management in high-altitude patients with elevated hemoglobin, and they cannot be invoked to conclude that no hemoglobin-targeted intervention is warranted in this population; if anything, the TRAIN signal cautions against extrapolating our null admission-hemoglobin finding to any hemoglobin-lowering strategy.

The association between surgical treatment and DCI (aOR 5.22) should not be misinterpreted as evidence that surgery causes ischemia. Treatment allocation in observational aSAH cohorts is subject to confounding by indication, and the apparent association is compounded by competing risk and time-dependent (immortal time) bias [31,32]: surgical candidates in this cohort presented with worse admission grades and heavier clot burdens, while the non-surgical group was enriched with patients who died or remained deeply comatose before the DCI window opened. Because DCI occurred at a median of 4.5 days while surgical patients had a median onset-to-surgery interval of 8 days, a proportion of DCI events likely preceded aneurysm securing, meaning that treating “ever operated” as a baseline covariate risks reverse causation, and patients who died before surgery had no opportunity to develop DCI. We could not fully resolve these biases: surgical treatment was analyzed as a time-fixed covariate, and complete death-timing data were unavailable, so neither a time-dependent Cox model nor a formal Fine–Gray competing-risk model could be fitted; a sensitivity analysis excluding the two very early deaths left the altitude estimate unchanged (Section 3.8), but the surgery–DCI association in particular remains vulnerable to residual confounding and immortal time bias. Recent meta-analytic evidence indicates that clipping is associated with higher DCI risk than coiling (pooled OR 1.57) [15]; importantly, that comparison is between two aneurysm-occlusion techniques and cannot be extrapolated to a contrast of surgical versus non-surgical management, and because we could not distinguish clipping from coiling, treatment modality represents an additional unmeasured confounder. The literature on surgical timing remains contradictory: surgery within the 3–10-day window was associated with DCI in a poor-grade cohort [33], whereas the larger and more extensively adjusted REDDISH cohort found no association between treatment delay and DCI [34], and a network meta-analysis suggests the timing–outcome relationship is etiology-specific and non-linear, with early occlusion framed primarily as rebleeding prevention [35,36]. Despite this uncertainty, one principle remains clear: timely aneurysm occlusion to prevent rebleeding—and to permit subsequent hemodynamic management—remains the cornerstone of aSAH care [1,37].

If replicated, residence above 4000 m could serve as an inexpensive, immediately available risk marker to flag patients warranting intensified surveillance during the 4–14-day DCI window, and to inform individualized decisions regarding inter-hospital transfer from ultra-high-altitude regions to lower-altitude referral centers before the vasospasm window—balanced against the risks of transport-related rebleeding and interrupted monitoring—consistent with contemporary guideline frameworks [1,37]. With respect to pharmacological prophylaxis, the appropriate inference from our data is not that nimodipine should be prolonged beyond the standard course, but rather that all patients should complete the guideline-recommended full 21-day course of enteral nimodipine [1,37]. The non-DCI group received a median of only 17 days of nimodipine, which may reflect truncation by early death or discharge, discontinuation after transfer, or under-prescription—each explanation carrying different implications for the quality of DCI prophylaxis at this center. No randomized trial has demonstrated benefit from extending nimodipine beyond 21 days, so our data support ensuring completion of the standard course rather than prolonging it. More broadly, our findings argue for developing altitude-adapted DCI prevention and monitoring protocols rather than uncritically extrapolating sea-level pathways; among candidate adjunctive strategies, prophylactic lumbar cerebrospinal fluid drainage reduced secondary infarction burden and improved six-month functional outcome in the randomized EARLYDRAIN trial [38], and represents a pragmatic, low-cost intervention that could be evaluated as part of an altitude-adapted DCI-prevention bundle in future prospective studies. Equally important is a methodological point: this study illustrates the value of explicit adherence to the 2010 consensus DCI definition, which remains inconsistently applied in the literature; standardized definitions are a prerequisite for pooling and meta-analyzing future high-altitude cohorts [3].

Several limitations should be considered when interpreting these findings. First, the single-center retrospective design and the referral-based, predominantly good-grade cohort (in-hospital mortality 7.4%) limit causal inference and generalizability to sea-level populations or to other high-altitude settings where erythrocytosis is more pronounced; these are constraints common to observational studies of this kind, and the consistency of the altitude effect across multiple sensitivity analyses provides some reassurance against chance or single-source bias. Second, residential altitude was used as a surrogate for chronic hypoxic exposure and ethnicity was not recorded, so the same altitude may correspond to different physiological doses in native Tibetans and Han migrants; accordingly, the null hemoglobin finding should be interpreted within this predominantly altitude-adapted population rather than extrapolated to high-erythrocytosis groups. Third, DCI was adjudicated retrospectively without full blinding or formal inter-rater reliability, and 25.9% of timed events fell outside the canonical 4–14-day window; although the consensus definition was applied and the altitude estimate remained stable (aOR 1.08–1.10) when the window was restricted, some early events may reflect early brain injury rather than true DCI. Fourth, death as a competing event and immortal time bias could not be fully modeled because death-timing data were incomplete, so the surgery–DCI association in particular should be regarded as exploratory; sensitivity analyses excluding very early deaths nonetheless left the altitude estimate unchanged. Fifth, several treatment and neurocritical-care variables (occlusion technique, CSF diversion, vasospasm monitoring, transfusion) and hemoglobin dynamics were not captured, and internal validation without an external cohort means the model should be viewed as etiological rather than as a validated prediction tool.

In conclusion, among high-altitude patients with aSAH, DCI occurred in 23.4% and was strongly associated with in-hospital mortality and 1-year disability. Greater hemorrhage burden (Fisher grade) and surgical treatment were independently associated with DCI, while admission hemoglobin was not. The altitude association—moderate in magnitude, biologically plausible, stable in point estimate across DCI definitions, but borderline and statistically unstable across model specifications, and non-monotonic in stratified analyses—suggests, but does not establish, that chronic high-altitude residence may act as an environmental modifier of DCI risk, possibly in a threshold-type pattern above approximately 4000 m rather than as a linear dose–response relationship. Multicenter prospective studies with objective exposure ascertainment, formal competing-risk and non-linear modeling, and mechanistic monitoring are needed to confirm or refute this hypothesis before any clinical pathways are modified.

## Data Availability

All data produced in the present study are available upon reasonable request to the authors

## Founding

This research was supported by the Natural Science Foundation of the Tibet Autonomous Region, Project Number: XZ2024ZR-ZY004 (Z).

## Interests and conflicts

There is no conflict of interest among the authors of this article.

## Notes

### Competing Interest Statement

The authors have declared no competing interest.

### Author Declarations

The study protocol was approved by the Ethics Committee of the People’s Hospital of Tibet Autonomous Region (XZ2024115).

### Summary of Updates

Version note (v2): This revision supersedes the preliminary analysis posted as v1. In v1, delayed cerebral ischemia (DCI) was defined using a more restrictive, manually adjudicated criterion (26 events, 10.2%) and residential altitude was analyzed in quartiles. In the present version, all DCI events were re-adjudicated against the 2010 international consensus definition (Vergouwen et al.), yielding 60 events (23.4%, consistent with the ~30% incidence in current guidelines), and altitude was modeled as a continuous exposure. Multivariable analyses, bootstrap internal validation with calibration assessment, and prespecified sensitivity analyses across three consensus-based DCI definitions are now reported; the altitude effect estimate was stable across definitions (adjusted OR 1.08-1.10 per 100 m). The revised findings reflect improved outcome ascertainment, statistical power, and methodology rather than a change in the underlying cohort.

